# Safety Assessment of BNT162b2 Vaccine in Adolescents Aged 12-15 Years

**DOI:** 10.1101/2021.11.06.21266016

**Authors:** Jayendrakumar Patel, Bhavesh Bhavsar, Shalin Parikh, Nikunj Ladani

**Author notes:** **Corresponding Author:** Jayendrakumar Patel, CEO and Executive Director, Pyrrhic Pharma USA, LLC, 8 The Green, Dover, DE, USA - 19901., **Corresponding author’s email:**.

## Abstract

Present evidence of the safety of BNT162b2 Vaccine in adolescents of 12 to 15 years relies only on 2260 subjects involved in phase 3 study. Therefore, clinical and post-clinical safety assessment of BNT162b2 Vaccine in Age Group of 12 to 15 Years is urgently needed to make an accurate judgment for mass vaccination. A rapid systemic review was conducted to determine safety profile of BNT162b2 Vaccine in Age Group of 12 to 15 Years following the PRISMA guidelines. Published literature before August 15, 2021 were searched in PubMed, Scopus, Web of Science, Embase, the Cochrane library and MedRxiv, using the defined search words. 135 records found from 6 databases, of which 4 studies (2 studies from NEJM and 2 MMWR), total subject who received at least one dose of vaccine: 64969, were included in this systemic review per the inclusion criteria. The major events reported in clinical phase and post-authorisation observational studies are pain at injection site (local), fatigue (systemic), headache (systemic), chill (systemic), diarrhoea (systemic) and joint pain (systemic). Post-authorisation observational study (n = 62,709) reported about 50% lower major systemic events, specifically, fatigue, headache, chill, diarrhoea and join pain and about 25% lower major local event, specifically, pain at injection site, than phase 3 clinical study (n = 1,131). Our study suggest that higher adherence rate (>97 percent received second dose) in clinical phase 3 and significantly lower incident of major local and systemic events in post-authorisation observational study indicating that BNT162b2 vaccine has highly favourable safety profile.

## INTRODUCTION

As of September 23, 2021, the coronavirus disease 2019 (Covid-19) pandemic has resulted in more than 230 million infections across all age groups in more than 190 countries, as well as more than 4.7 million deaths worldwide.^**1**^ There are several options for effectively treating COVID 19 infection, with vaccine being the most promising strategy in combating the global COVID-19 pandemic.^**2**^ BNT162b2 (Pfizer–BioNTech) is a Covid-19 vaccine that contains nucleoside-modified messenger RNA encoding the spike glycoprotein of the severe acute respiratory syndrome coronavirus 2 (SARS-CoV-2), which is associated with severe acute respiratory syndrome.^**3**^ High neutralising titers and robust antigen-specific CD4+ and CD8+ T-cell responses against SARS-CoV-2 were elicited in healthy persons after two 30-μg doses of BNT162b2.^**4,5**^ When tested in the phase 2–3 part of an ongoing global, multicenter, randomised, controlled trial involving participants 16 years or older, BNT162b2 demonstrated a favourable safety profile, with transient mild–moderate injection site pain, fatigue, and headache as the most common side effects. It also demonstrated a 95 percent efficacy in preventing Covid-19 from 7 days after the second dose.^**6**^ As a result of these findings, the Food and Drug Administration granted an emergency use authorization of BNT162b2, for the prevention of Covid-19 in people 16 years of age and older, on December 11, 2020.^**7**^ On May 10, 2021, the emergency use authorisation (EUA) was expanded to include adolescents aged 12–15 years, based on the results of a Phase 3 clinical trial.^**8,9**^ Present evidence of the safety of BNT162b2 Vaccine in adolescents of 12 to 15 years in adolescents of 12 to 15 years relies only on 2260 subjects involved in phase 3 study.^**9**^ More adverse events following immunisation (AEFI) have been observed as mass vaccination has progressed, particularly rare AEFIs. As instance, cases of myocarditis after vaccinating with the Pfizer-BioNTech vaccine began to be reported in June 2021 after receiving the second dose, primarily among younger males.^**10,11**^ Although, after reviewing available data, the centre for disease control and prevention (CDC’s) Advisory Committee on Immunization Practices (ACIP) concluded on June 23, 2021 that the benefits of BNT162b2 Vaccine in adolescents of 12 to 15 years to the population outweighed the risks of myocarditis, therefore ACIP recommended the vaccine continued used in people aged 12 years and older.^**12**^ For public confidence and to enable rapid, evidence-based policy decisions for population-level usage, it is necessary to demonstrate and summarise vaccine safety from clinical trials and post-authorization surveillance. Therefore, we conducted a systematic review to determine and compare the safety of BNT162b2 (Pfizer–BioNTech) vaccine between clinical phase 3 study and post-authorization safety surveillance.

## METHODS

### Search Strategy and Study Selection

The Preferred Reporting Items for Systematic Reviews and Meta-Analyses (PRISMA) guidelines^**13**^ were followed in the conducting of our systematic review. Published literature before August 15, 2021 were searched by J.P. in PubMed, Scopus, Web of Science, Embase and the Cochrane library, using the search words “(Pfizer-BioNTech COVID-19 Vaccine) and (BNT162b2) and (Pfizer–BioNTech) and (SARS-CoV-2 or COVID-19 or 2019–nCoV) and (Vaccine) and (12 - 15 years) and (Adverse Events or Safety) and (Efficacy)”. In addition, we searched the preprint database MedRxiv for any publications that might be connected to our research. The search was restricted to papers written in the English language only.

Only manuscripts that published in a scientific peer-reviewed journals, preprints published in MedRxiv and unpublished data disclosed by only regulatory or public health agencies, i.e., the United States Food and Drug Administration (USFDA) and the Centers for Disease Control and Prevention (CDC), were included in our search for clinical trials and post-authorization observational studies that examined safety data of BNT162b2 Vaccine in Age Group of 12 to 15 Years. Exclusion: We excluded media news, commentaries, study protocols, case reports, reviews, and scientific conference abstracts as well as manuscripts published in scientific journals or interim reports or final reports of clinical trials those not explicitly disclosing safety data of BNT162b2 Vaccine in Age Group of 12 to 15 Years. For post-authorization observational studies with sample sizes of less than ten thousand participants were also excluded.

### Data Extraction and Quality Assessment

Three researchers (J.P., B.B., and S.P.) evaluated eligible studies, did data extraction, and cross-checked their findings to ensure accuracy. Second and third researchers, B.B. and S.P. respectively, independently extracted data and any discrepancies in the data were resolved through discussion with J.P., a first researcher. Data extracted from the qualified studies included name of first author, time of publication, study details (e.g. clinical phase and post-authorisation observational study), characteristics of subjects (age group), intervention measures (number of doses administered, i.e., dose 1 and dose 2, etc.), the number of subjects in the safety dataset, and the rate of AEFI during the follow-up period. Where some require data not available for concluding present study were calculated from the reported data in the study accordingly. If data for the same subjects or study were presented in multiple publications, these data were only counted once. Due to phase 1 and 2 trials often including multiple differing experimental groups (different age group), we focused exclusively on safety data from experimental groups of age 12 to 15 years in phase 3 clinical trials. Certainty of evidence was assessed by researchers according to the grading recommendations assessment, development and evaluation (GRADE) framework. ^**14,15**^

### Data synthesis and Analysis

For the safety profile of BNT162b2 Vaccine in Age Group of 12 to 15 Years in phase 3 clinical trials, the primary outcomes were the proportion of vaccine recipients experiencing at least one AEFI and the rates of selected AEFI of BNT162b2 Vaccine versus placebos. In the BNT162b2 Vaccine immunization strategies, at least two doses are usually needed to achieve an ideal protection efficacy, and we, therefore, also compared the AEFI incidence between the priming dose and the booster dose (dose 1 vs. dose 2).

For post-authorization observational safety data of dose 1 vs dose 2 of BNT162b2 Vaccine, we examined rates of AEFIs, local adverse events, systemic adverse events and any health impact.

As phase 3 clinical included only small portion of subjects (2260), we also compared adverse events [local adverse events (e.g., pain at injection site, redness and swelling) and systemic adverse events (e.g., fever, fatigue, headache, chills, vomiting, diarrhoea, muscle pain and joint pain)] post-dose 1 and dose 2 of BNT162b2 Vaccine between phase 3 clinical studies Vs Post clinical observational studies to assess the trends of AEFIs of BNT162b2 Vaccine.

## RESULTS

### Study Inclusion and its Characteristics

The flowchart of literature screening in this study is shown in Figure 1. In our preliminary search, we got 135 records from 6 databases. According to predefined criteria, 4 studies (total subject who received at least one dose of vaccine: 64969) were finally included in this systemic review.^**9,16,17,18**^ Of these, 2 studies were officially published in The New England Journal of Medicine (NEJM), and 2 studies were published by CDC in Morbidity and Mortality Weekly Report. The main characteristics of included studies reporting safety of BNT162b2 Covid-19 vaccine in adolescents are reported in table 1.

**Table 1:**
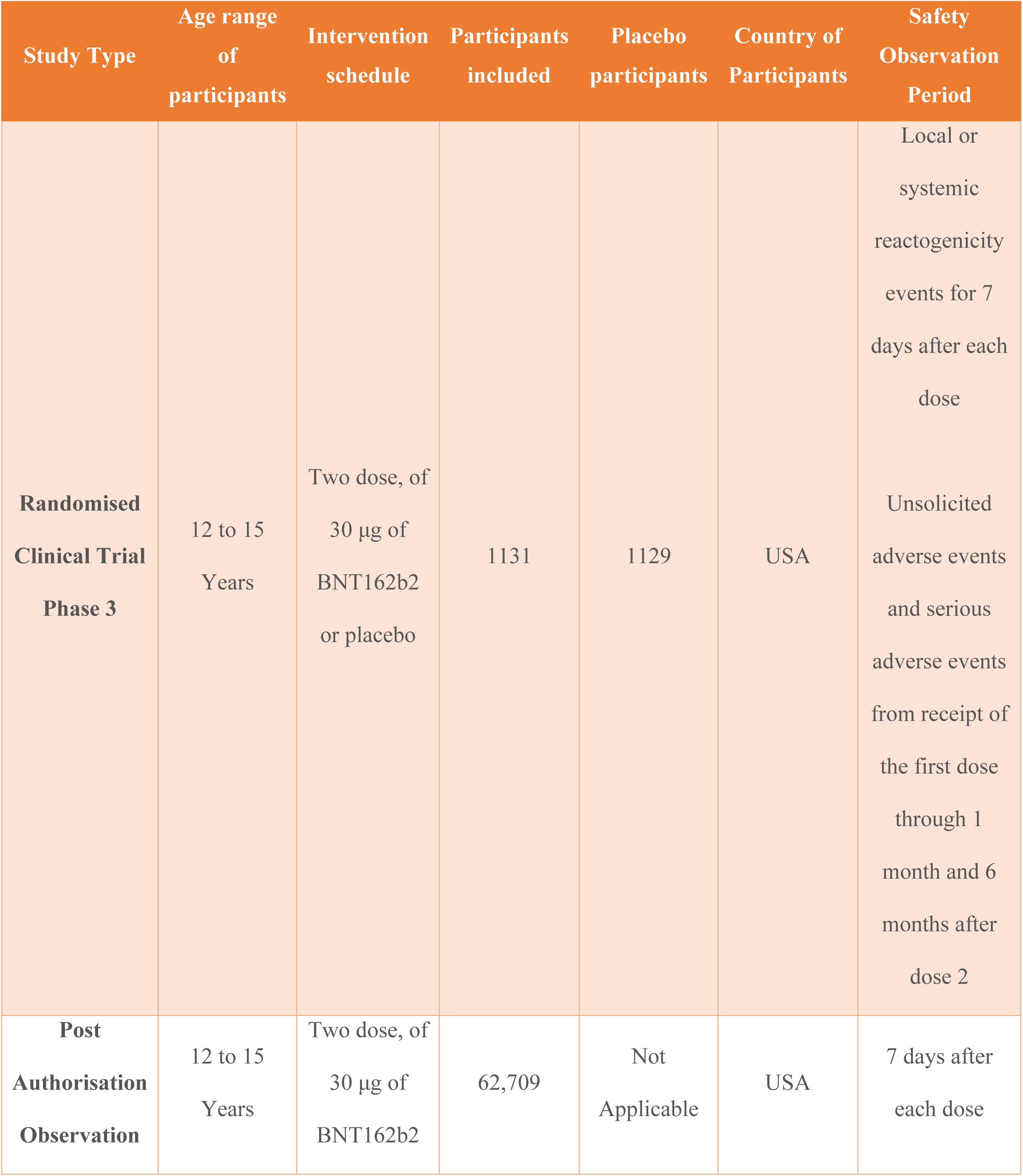
Characteristics of included studies reporting safety of BNT162b2 Covid-19 vaccine in Adolescents.

**Figure 1:**
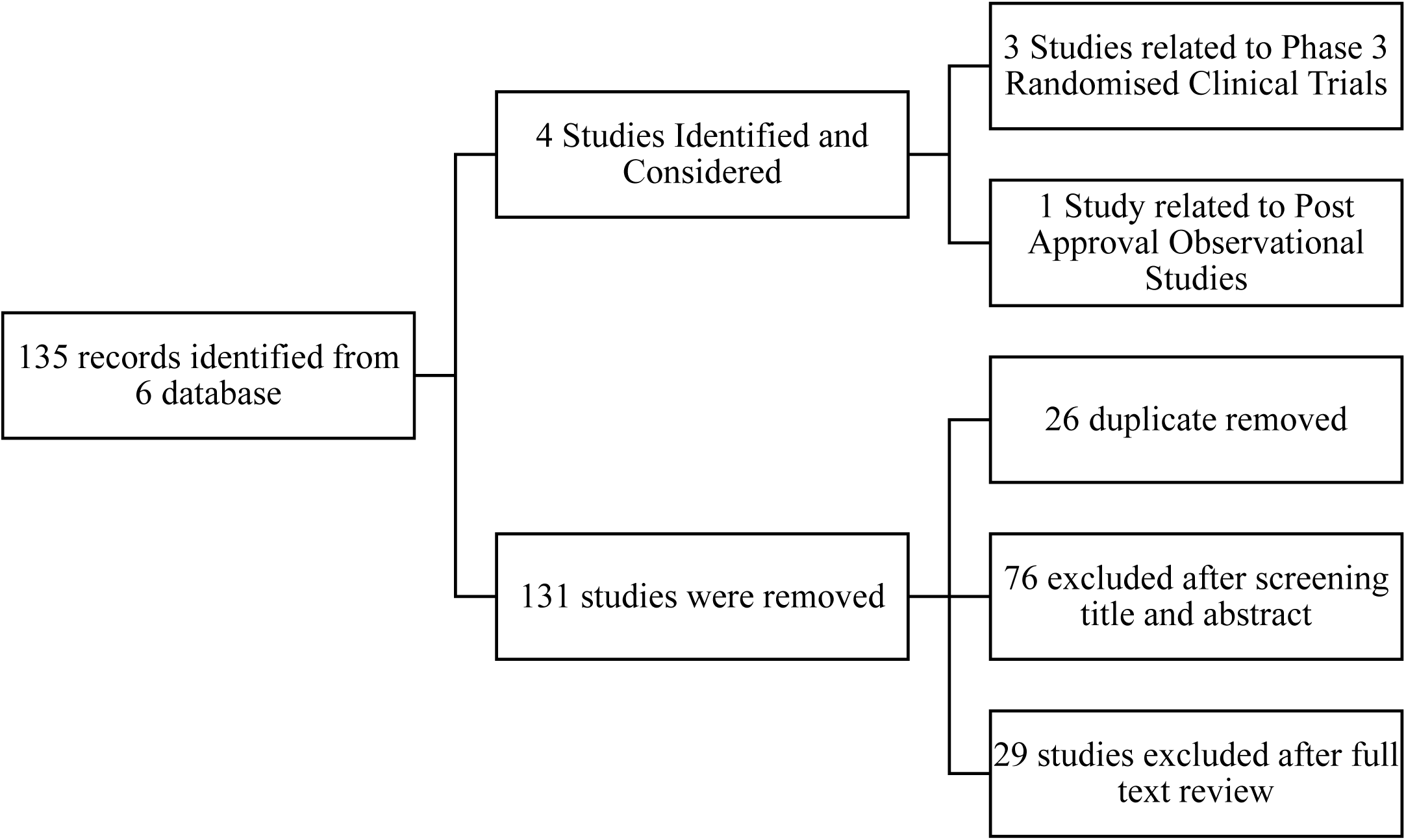
**The flowchart of Studies screening from 6 different database**

### Safety Assessment of Randomised Clinical Trails (Phase 3)

In total, 2260 adolescents between the ages of 12 and 15 years old received first dose intramuscular injections [1131 received 30μg of BNT162b2 vaccine and 1129 received 30μg of placebo (saline)], and 2241 adolescents between the ages of 12 and 15 years old received second dose intramuscular injections [1124 received 30μg of BNT162b2 vaccine and 1117 received 30μg of placebo (Saline).^**9,18**^ Following each of the two doses [figure 2 (A)], the most frequently reported reactions (figure 2) were injection-site pain in 86% of participants post-dose 1 vs 79% of participants post-dose 2, fatigue in 60% of participants post-dose 1 vs 66% of participants post-dose 2, headache in 55% of participants post-dose 1 vs 65% of participants post-dose 2, chills in 28% of participants post-dose 1 vs 42% of participants post-dose 2 and muscle pain (myalgia) in 24% of participants post-dose 1 vs 32% of participants post-dose 2.^**9,18**^

**Figure 2:**
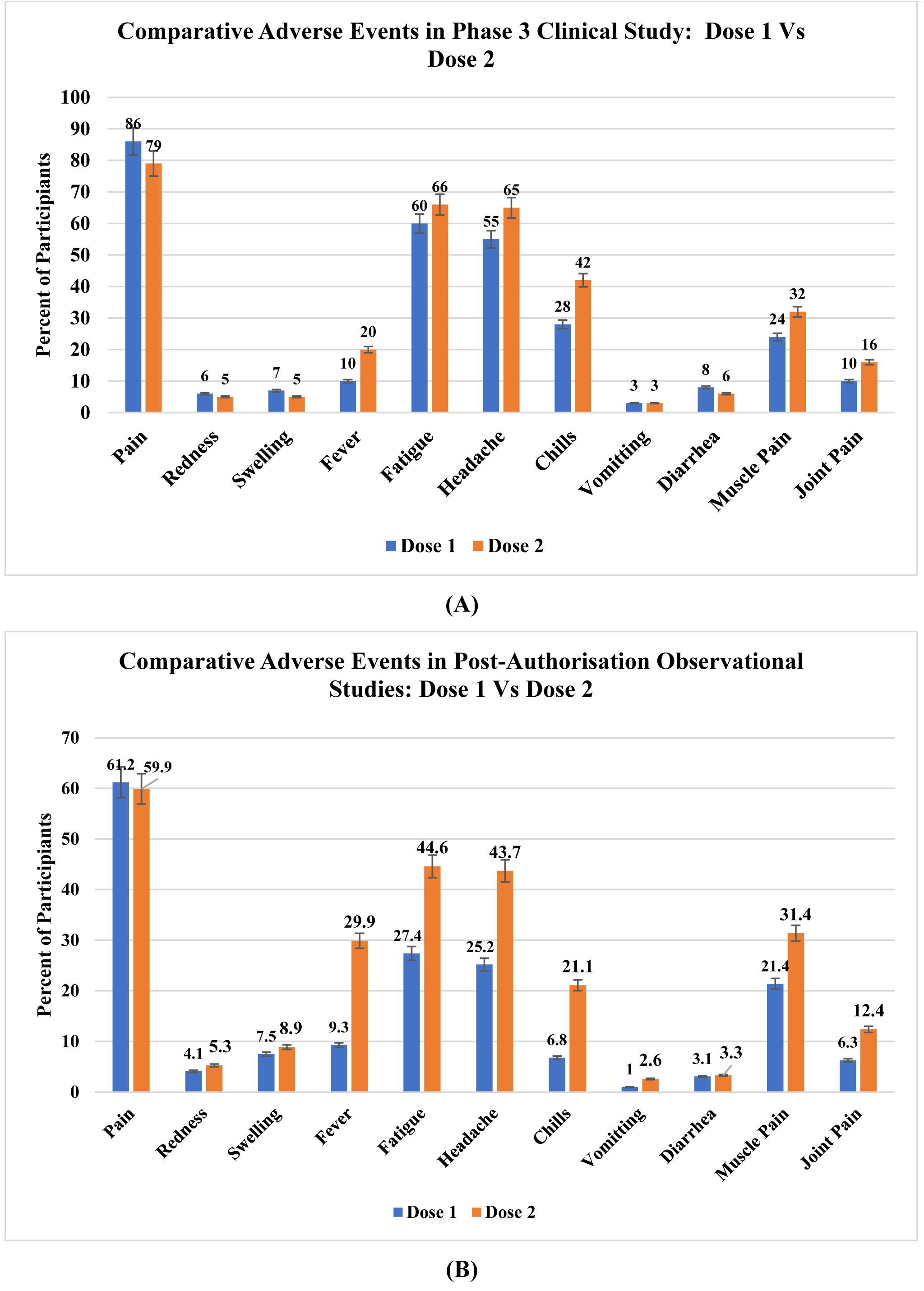
**(A) Comparative Adverse Events in Phase 3 Clinical Study: Dose 1 Vs Dose 2, and (B) Comparative Adverse Events in Post-Authorization Observational Studies: Dose 1 Vs Dose 2**

Overall, those who received BNT162b2 vaccine reported more local and systemic incidents than those who received placebo (saline). In most cases, local and systemic events were of mild to moderate severity, and they were usually resolved within 1 or 2 days of occurrence. The most often reported local reaction was pain at the injection site (86% post-dose 1 vs 79% post-dose 2), of them 1.5 percent participants reported severe injection-site pain after receiving any BNT162b2 dose, whereas no severe pain was reported after receiving a placebo. In case of systemic events, headache, fatigue, chills, fever and muscle pain were the most frequently reported. 20% participants reported fever (oral body temperature, ≥38°C) post-dose 2 compared to 10% participant post-dose 1, wherein one participant reported body temperature > 40°C one day after receiving dose 1 who was then discontinued vaccination although fever resolved after 2 days.^**9**^

Post-dose 1 through 1 month after dose 2, about 6% participants of both, BNT162b2 and placebo participants, were reported any events, of which 0.4% participants of BNT162b2 vaccine and 0.1% participants of placebo were reported any serious adverse events. Lymphadenopathy was reported in 9 of 1131 BNT162b2 recipients (0.8%) and in 2 of 1129 placebo recipients (0.2%). There were no thromboses, hypersensitivity adverse events, or vaccine-related anaphylaxis observed in this study. There were no reported fatalities.^**9**^

In general, local and systemic effects were reported more frequently after dose 2 of BNT162b2 vaccine than after dose 1.

### Post-Authorisation Observational Study

The V-safe system of the US Centers for Disease Control and Prevention (CDC) enrolled 62,709 adolescents beginning May 10, 2021 for safety observational study. During the week after receipt of dose 1, local (63.9%) and systemic (48.9%) reactions were commonly reported by adolescents aged 12–15 years; systemic reactions were more common after dose 2 (63.4%) than dose 1 (48.9%).^**17**^

Following each of the two doses [figure 2 (B)], the most frequently reported reactions were injection-site pain in 61% of participants post-dose 1 vs 60% of participants post-dose 2, fatigue in 27% of participants post-dose 1 vs 45% of participants post-dose 2, headache in 25% of participants post-dose 1 vs 44% of participants post-dose 2, Chills in 7% of participants post-dose 1 vs 21% of participants post-dose 2, muscle pain (myalgia) in 21% of participants post-dose 1 vs 31% of participants post-dose 2 and fever in 9% of participants post-dose 1 vs 30% of participants post-dose 2. Post-dose 2 of vaccination, about 23% participants reported that they are unable to perform normal daily activities and about 6% participants reported that they are unable to attend the school (figure 3). For each dose, reactions were reported most frequently the day after vaccination. ^**17**^

**Figure 3:**
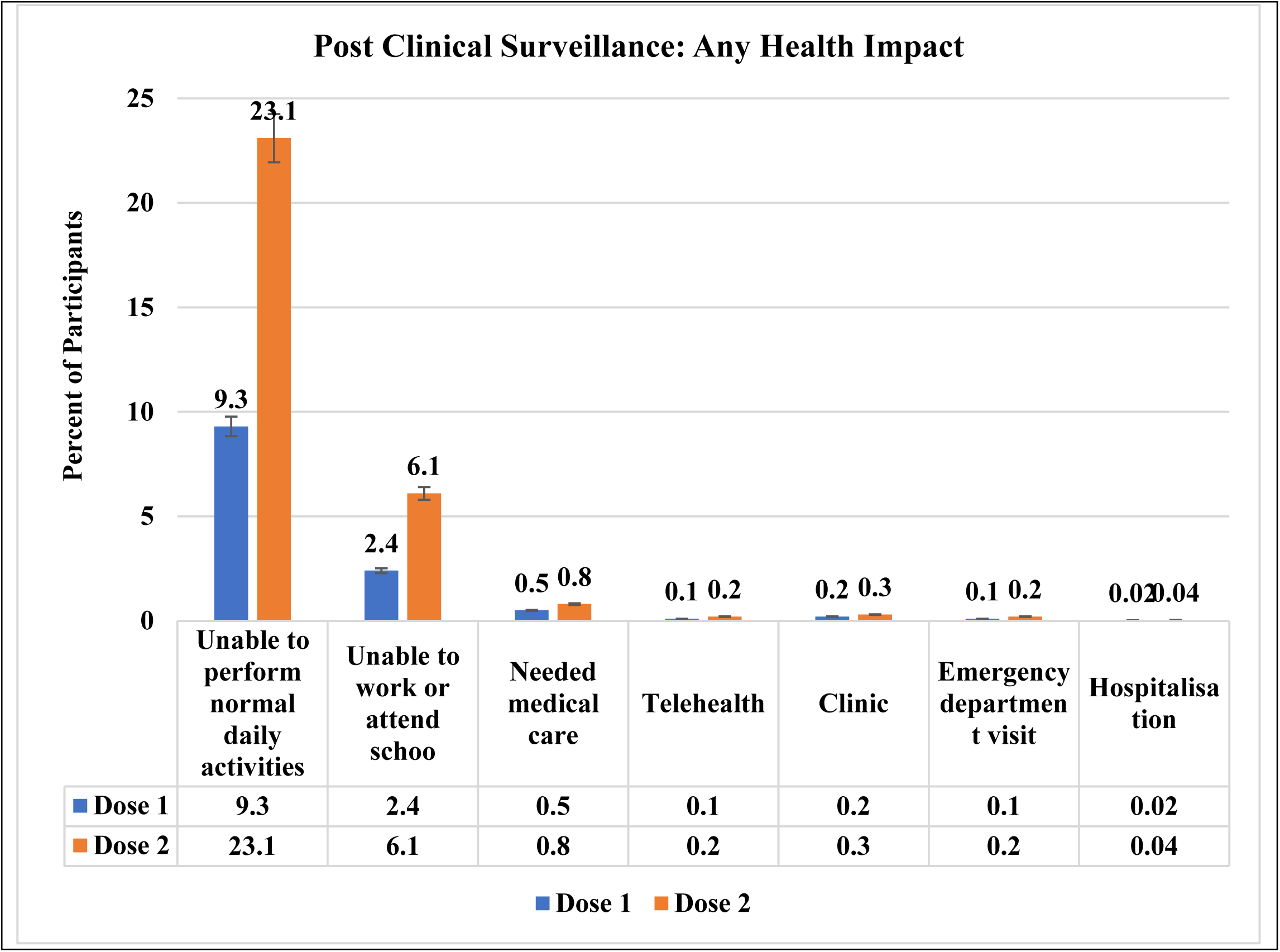
**Post Clinical Surveillance: Any Health Impact**

## DISCUSSION

An investigational two-dose regimen of 30μg BNT162b2 vaccine provided at 21 days interval to adolescents 12 to 15 years of age was found immunogenic, and the observed vaccination effectiveness against Covid-19 was 100 percent from 7 days after the second dose in phase 3 clinical study. The similar favourable safety profile was seen in adolescents 12 to 15 years of age who received two-dose regimen of 30 μg BNT162b2 vaccine as adults in the pivotal study and through continuous pharmacovigilance following vaccine approval. More than 97 percent of 12 to 15 years adolescents’ participants received second dose of BNT162b2 vaccine within the recommended time period indicating that adherence was very high.^**9, 18**^

The most often reported local reaction was pain at the injection site. In case of systemic events, headache, fatigue, chills, fever and muscle pain were the most frequently reported. Systemic events reported more often post second dose of the vaccine than dose 1, which is in line with previously reported trend of systemic events in age group of 16 years or older.

Phase 3 clinical safety data of BNT162b2 vaccine are in decline trend compared to the safety data of post-authorisation observational study, except very few events of myocarditis, a serious adverse event reported in post authorisation observation study. In phase 3, a total of 1,131 individuals aged 12–15 years who received vaccine reported local and systemic reactions that were largely mild (i.e., did not interfere with activity) or moderate (some interference with activity) and they were usually resolved within 1 or 2 days of occurrence; and no significant adverse events associated with vaccine were reported. In post-authorisation observational study, a total of 62,709 individuals aged 12-15 years who received the vaccine reported similar types of local and systemic events as like Phase 3 recipients, about 23% participants reported that they are unable to perform normal daily activities and about 6% participants reported that they are unable to attend the school. ^**17**^ The major events reported in clinical phase and post-authorisation observational studies are pain at injection site (Local), fatigue (systemic), headache (systemic), chill (systemic), diarrhoea (systemic) and joint pain (systemic). Post-authorisation observational study (n = 62,709) reported [figure 4 (A), (B), (C), (D)] about 50% lower major systemic events, specifically, fatigue, headache, chill, diarrhoea and joint pain and about 25% lower major local event, specifically, pain at injection site, than phase 3 clinical study (n = 1,131), whereas rest systemic events and local events are almost consistent between the clinical study and post-authorisation observational study. ^**17**^ Higher adherence rate (more than 97 percent participants received second dose within the recommended time period) in clinical phase 3 and significantly lower incident of major local and systemic events in post-authorisation observational study indicating that BNT162b2 vaccine has highly favourable safety profile in adolescents aged 12-15 Years.

**Figure 4:**
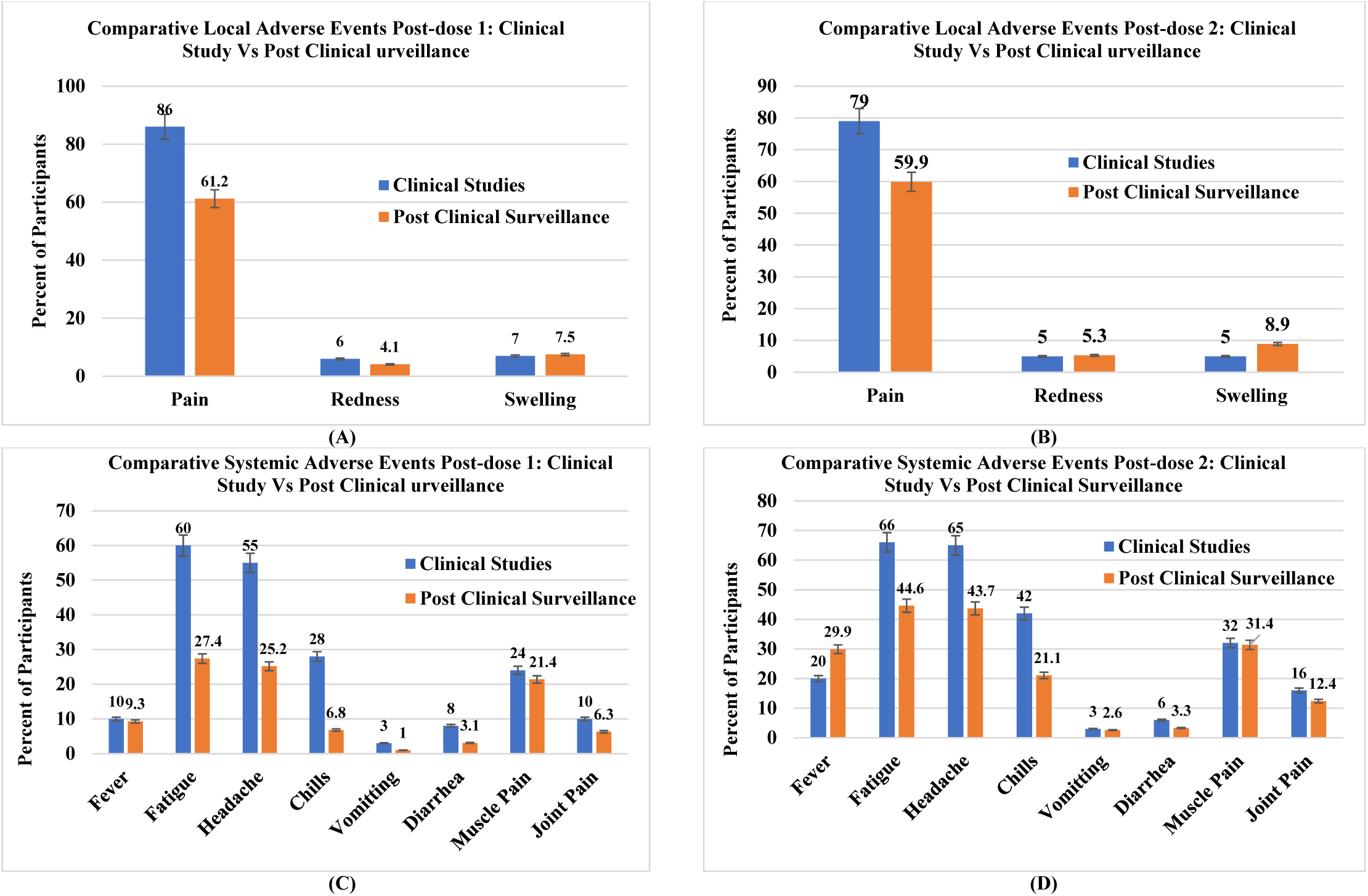
**(A) – Comparative Local Adverse Events Post-dose 1: Clinical Study Vs Post Clinical urveillance (B) – Comparative Local Adverse Events Post-dose 2: Clinical Study Vs Post Clinical urveillance (C) – Comparative Systemic Adverse Events Post-dose 1: Clinical Study Vs Post Clinical urveillance (D) - Comparative Systemic Adverse Events Post-dose 2: Clinical Study Vs Post Clinical Surveillance**

Our research has a few limitations: First, our findings are based on very limited studies because the EUA for the BNT162b2 Vaccine in the age group of 12 to 15 years has been extended on May 10, 2021. Second, this study includes only short-term safety data for the BNT162b2 vaccine. Long-term safety data for the BNT162b2 vaccine in the age group of 12 to 15 years are not available. Third, v-safe safety data are taken into account for post-authorization observational studies. v-safe is a voluntary self-enrolment programme promoted by vaccine administrators that requires children aged below 15 years to be enrolled by a parent or guardian. As a result, v-safe data has some limitation to be generalizable to the entire population of vaccinated adolescents.

## Data Availability

All data produced in the present work are contained in the manuscript.

## AUTHORSHIP

**Jayendrakumar Patel:** Conceptualisation of research design and research method. Data curation, formal analysis and validation of data, investigation, writing first original draft manuscript and editing. **Bhavesh Bhavsar and Shalin Parikh:** These authors contributed equally in Data curation and formal analysis. Review first original draft manuscript.

## ETHICAL APPROVAL

Not Applicable

